# Whole-brain structural and functional neuroimaging of individuals who attempted suicide and people who did not: a systematic review, exploratory coordinate-based meta-analysis and seed-based connectivity study

**DOI:** 10.1101/2022.10.21.22281350

**Authors:** Nicola Meda, Alessandro Miola, Giulia Cattarinussi, Fabio Sambataro

## Abstract

**Introduction:** Suicide is the cause of death of approximately 800’000 people a year. Despite the relevance of this behaviour, risk assessment tools rely on clinician experience and subjective ratings.

**Methods:** Given that previous suicide attempts are the single strongest predictors of future attempts, we designed a systematic review and coordinate-based meta-analysis to evince if any neuroimaging features can help distinguish individuals who attempted suicide from subjects who did not. Out of 5659 publications from PubMed, Scopus and Web of Science, we summarised 102 experiments and meta-analysed 23 of them.

**Results:** A cluster in the right Superior Temporal Gyrus, a region implicated in emotional processing, might be functionally hyperactive in individuals who attempted suicide. Furthermore, we used JuSpace and the Human Brain Connectome dataset to show that this cluster is enriched in 5-HT_1A_ heteroreceptors, and its connectivity with the left central operculum is significantly correlated with loneliness scores.

**Conclusions:** This evidence provides a putative neural substrate for suicidal thought-to-attempt transition as hypothesised by Joiner’s Interpersonal Theory of Suicide. Heterogeneity in the analytical techniques and weak or absent power analysis of the studies included in this review currently limit the applicability of the findings, the replication of which should be prioritised.

## Introduction

Suicide is a behaviour that leads to the premature death of approximately 800’000 people a year (*WHO* | *Suicide data*, 2020). It is estimated that for each death by suicide there are approximately 10 to 25 non-fatal suicide attempts (SAs) (Stone et al., 2021; Wasserman, 2016). However, the SA to death ratio might be higher in specific age ranges, like adolescence (Maris, 2002). In light of the epidemiological relevance of suicidal behaviours, the importance of recognising people who are at risk of death by suicide cannot be overstated. Despite the enormous amount of published literature on lifetime (Haney et al., 2012; Masango et al., 2008; Mościcki, 1997) and proximate risk factors (Berman, 2018) of suicide, we are nowhere close to predicting suicidal behaviour with accuracy, to the extent that some authors stated that “suicide is easier to prevent than to predict” (Rihmer et al., 2018), at least for specific groups of people. Among the myriad of factors associated with an increased risk of death by suicide, the most relevant is previous SAs (Christiansen & Frank Jensen, 2007; Owens et al., 2002). A recent systematic review and meta-analysis pooling data from 41 previous reports estimated that the death by suicide rate sits at 2.8% (2.2-3.5) one year, 5.6% (3.9-7.9) five years, and 7.4% (5.2-10.4) ten years after a nonfatal suicide attempt (Demesmaeker et al., 2022). Indeed, people who acted on their thoughts of death once are more likely to further attempt, as this dramatic solution to the mental pain they experienced has become cognitively more accessible (R. C. O’Connor & Portzky, 2018), and the bridge between ideation and action has already been crossed once (Ken Norton, 2018; R. O’Connor, 2021). There are several factors, or “bridges”, that make the ideation to action transition more likely (O’Connor & Kirtley, 2018), that can be grouped into four clusters, and are typically investigated to assess the suicide risk of an individual: past experiences (previous (own) SAs and exposure to (family member/friends) suicidal behaviour); acquired capability (Chu et al., 2017; Joiner & Jr, 2005) (encompassing fearlessness about death and increased physical pain endurance); access to means and plans for suicide (Jobes, 2016; Stanley & Brown, 2012); impulsivity (Gvion & Apter, 2011). In our opinion, mental imagery could be grouped with past experiences (if the individual visualisation of dying is influenced by previous attempts or exposure), access to means and plans (when the visualisation of dying or death relates to a plan) or it might pertain to intrusive thoughts. The previous factors are pivotal in ascertaining if an individual is at risk of death. However, they can be identified only through a thorough suicidal risk assessment. Mostly, this procedure is subjective and dependent on the patient’s expected consequences of their statements and the patient-interviewer therapeutic alliance. Thus, it is of utmost importance to investigate if any objective finding can aid the interviewer in distinguishing which patients are more likely to attempt suicide. One step in this direction could be taken by examining if any neuroimaging findings can distinguish the people who attempted suicide from those who did not. Then, if any difference emerged, it would be easier to test prospectively if said diversity can be leveraged to aid the suicidal risk assessment. Brain imaging and connectivity data had been used to train and test predictive models that could identify people with or without suicidal ideation (Stumps et al., 2021); Previous studies also showed that brain regions implicated in decision-making (e.g., frontal areas) or emotion regulation (e.g., temporal cortex, insula) might help discern people with a history of suicidal behaviour from those without it with good accuracy (Aguilar et al., 2008; Stange et al., 2020; Zhu et al., 2020), thus indicating that an imaging-based approach could be leveraged to increase the accuracy of suicidal risk assessment. One aspect that is as crucial as frequently overlooked is that any diversity between people who attempted suicide and people who did not should be transdiagnostic, i.e., shared across the different mental health conditions that the individual might be suffering from. The reasons are twofold: although mental disorders are known risk factors for death by suicide (Moitra et al., 2021), mental health conditions are not a prerequisite nor sufficient to explain suicidal behaviours. Most people who die by suicide have no history of any mental health disorders (Too et al., 2019). Moreover, a large proportion of people who died had no mental health condition identifiable through psychological autopsy (Cavanagh et al., 2003). A similar picture emerges when considering the individuals who survived the first SA (Nock et al., 2009). Secondly, the suicidal risk assessment usually takes place at first contact, so this usually happens well before reaching a diagnosis of mental disorder. Most research on the neuroimaging features of suicidal behaviours has been conducted on participants who suffered from mood disorders (Campos et al., 2021; Duarte et al., 2017; S. Fan et al., 2019; for the ENIGMA-Major Depressive Disorder Working Group et al., 2017; Johnston et al., 2017; S.-G. Kang et al., 2020; Lee et al., 2016; Zhu et al., 2020). However, it was suggested that some brain regions, such as the temporal lobe, might be shared hubs of dysfunction associated with suicidal behaviours across different mental health conditions (Domínguez-Baleón et al., 2018). To our knowledge, this hypothesis has not been meta-analytically investigated so far. Two previously published reviews addressed the issue of common features of suicidality across mental health conditions but either included non-suicidal self-harm in the analysis (X. Huang et al., 2020) or included studies that did not look for brain-wise differences (Jollant et al., 2018) but that instead focused on specific areas of the brain (Soloff et al., 2012) thus producing a sampling bias.

Therefore, we conducted this systematic review and meta-analysis to summarise the published literature to explore whether people with a history of SA have structural or functional brain differences with respect to people without such history (NSA). Subsequently, we performed spatial correlation analysis with neurotransmitter receptor maps (based on PET scans) to evince if any of the meta-analytically identified brain regions is significantly associated with a particular receptor expression. Lastly, we applied seed-based functional connectivity (FC) analysis to a sample of healthy subjects to investigate the resting-state FC of the brain areas identified from the meta-analysis of resting-state fMRI studies.

## Methods

### Protocol and search strategy

This systematic review and coordinate-based meta-analysis followed a pre-defined protocol published at (https://osf.io/fvrdx) and adhered to the procedures of the Preferred Reporting Items for Systematic Reviews and Meta-Analyses (PRISMA) statement (Page et al., 2021)) (see Supplementary materials for details and PRISMA Checklist (Title, 2009)). A comprehensive literature search was performed in PubMed, Scopus, and Web of Science databases, with the following keywords: ((“suicide” OR “suicidal” OR “suicidality” OR “self-harm” OR “self-injury” OR “self-directed” OR “self-mutilation”) AND (“MRI” OR “magnetic resonance” OR “magnetic resonance imaging” OR “brain imaging” OR “neuroimaging” OR “functional MRI” OR “fMRI” OR “functional magnetic resonance imaging”)) from inception until May 19, 2022. A manual search was conducted on the reference list of the included studies, relevant review articles, International Clinical Trials Registry Platform. Commentaries, editorials, and reviews were excluded. No language or publication status restriction was applied.

### Eligibility

Experimental, cross-sectional, case-control and prospective magnetic resonance imaging (MRI) studies were considered eligible. Studies were included if 1) they reported the results of structural, or resting-state, or task-based functional MRI, or FC, or cortical thickness (CT) of SA and NSA (aged > 18 years); 2) the two groups of people being compared were homogeneous in terms of mental health condition (MHC; i.e., people with schizophrenia (SCZ) that attempted suicide (+AS) were compared to people with SCZ who did not attempt suicide at the time of scanning). In the case that a study reported two heterogeneous groups (i.e., people with SCZ + AS compared with otherwise healthy controls), the study was excluded; 3) the primary mental health condition had to be assessed with a semi-structured or structured interview and diagnosed according to the criteria listed in the Diagnostic and Statistical Manual (DSM) or the International Classification of Diseases (ICD); 4) the structural and/or functional studies reported the stereotaxic coordinates of the significant clusters of MHC+AS vs MHC-AS as well as full details on imaging acquisition to allow an unequivocal assessment of the extent of brain coverage. In particular, the field of view (or the matrix and voxel volume), slice number, and slice thickness had to be reported. If the product of (slice thickness + gap) times slice number was less than 93mm (the average axial diameter – height – of the brain excluding the cerebellum), the study probably did not cover the whole brain. It thus was flagged and considered together with studies with similar brain coverage. If the product was less than the average brain + cerebellum height (average 105mm), the study was regarded as likely covering the entire brain except for the cerebellum. Nonetheless, it was considered and eventually meta-analysed with other studies with overlapping brain coverage. In the case that a study acquired images of the entire brain (including the cerebellum) but did not consider the cerebellum or other volumes in the statistical analysis, it was regarded as “brain coverage only” (i.e., excluding the cerebellum) or partial brain coverage (i.e., if it excluded the occipital lobe). A statement that the “whole brain” was covered was deemed insufficient, and NM proceeded to contact the authors of the study, requesting further details. In the case that NM did not receive a reply in two weeks (after a second solicitation), the study was excluded from the meta-analysis; 5) studies that regarded *a priori*-defined regions-of-interest (ROI) were excluded. Studies that applied small volume corrections (SVC) were excluded unless there was the possibility to include those clusters that reached statistical significance with whole-brain-level correction; 6) studies on task-based functional neuroimaging were grouped according to the task administered or, in case of high heterogeneity of tasks, on the cognitive domain investigated; 7) studies regarding CT were analysed separately from the structural and functional studies; 8) studies regarding seed-based FC were grouped according to seed localisation. Studies that investigated FC with Independent Component Analysis (ICA) were separated from the studies employing a seed-based approach. In general, the following guidelines (Müller et al., 2018; Tahmasian et al., 2019) for conducting a neuroimaging meta-analysis were followed.

### Data extraction

Duplicate records were excluded. Two authors (NM, AM) independently extracted each eligible study characteristics and outcome data. Any disagreement was solved by a third author (FS). The following variables were extracted: DOI, first author, year of publication, study design, country where the research took place, primary diagnosis of the participants, DSM version used for psychiatric assessment, sample size per group, demographics (sex, age, education, illness duration), neuroimaging analysis methods (e.g., for structural studies: VBM; resting-state index studies: amplitude of low-frequency fluctuations (ALFF) (static, dynamic, normalised); for FC: seed localisation or if the authors employed ICA; for task-based investigations: task characteristics and cognitive domain), medication, comorbid substance abuse (e.g., alcohol, drugs), weeks since last SA to scan, rating scales (e.g., Hamilton Depression Rating Scale, Beck Depression Inventory-II or others), scanner magnetic field strength, software used for the analysis (e.g., SPM5, SPM8), stereotaxic coordinate system (MNI, Talairach), the direction of contrast, coordinates, statistical correction. Studies that did not report any significant differences in brain volumes between the two groups (i.e., that could not pinpoint any coordinates that differentiated people with SA from NSA) could not be included in the coordinate-based meta-analysis due to methodological reasons explained elsewhere (Müller et al., 2018; Tahmasian et al., 2019). However, these studies are acknowledged in Supplementary Appendix.

### Data analysis

A tailored version (see Supplementary Appendix and (Cattarinussi et al., 2019)) of the Imaging Methodology Quality Assessment Checklist (Shepherd et al., 2012) was used to assess a quality score for each study included in the coordinate-based meta-analysis (reported in Table 1), and both reviewers (NM, AM) independently assessed each study for the risk of bias.

**Table 1.** Characteristics of the studies included in the coordinate-based meta-analysis and quality score. BD = Bipolar Disorder; CBMA = coordinate-based meta-analysis; CT = Cortical Thickness; DSM = Diagnostic and Statistical Manual of Mental Disorders; FC = functional connectivity; FEP = First-episode psychosis; FEW = Family-wise error; GMV = grey matter volume; MDD = Major Depressive Disorder; MHC = mental health condition; NSA = Non-suicide attempt; rs-fMRI = resting-state functional magnetic resonance imaging; SA = Suicide attempts; SCZ = Schizophrenia; VBM = Voxel-based morphometry.

| Type of analysis | Study | N (SA/NSA) | Age (mean±sd, SA/NSA) | Female % (SA/NSA) | MHC | Diagnostic Criteria | Time from SA to scan | Quality Score |
| --- | --- | --- | --- | --- | --- | --- | --- | --- |
| VBM | Wang et al., 2020 | 70/128 | 27.5±9.6/<br>27.1±8.3 | 78.5/ 72.6 | BD or MDD | DSM-IV | NA | 8 |
|  | Peng et al., 2014 | 20/18 | 27.7±7.2/<br>31±7.4 | 65/66 | MDD | DSM-IV | NA | 8.5 |
|  | Johnston et al., 2017 | 26/42 | 20.5±3/<br>20.6±3.2 | 77/55 | BD | DSM-IV | NA | 9.5 |
|  | Lee et al., 2016 | 19/19 | 41.9±10.8/<br>41.1±15.1 | 57.9/47.4 | MDD | DSM-IV | NA | 9 |
|  | Aguilar et al., 2008 | 13/24 | 37.1±10/<br>42.6±10.1 | 0 | SCZ | DSM-IV | NA | 8 |
|  | Jollant et al., 2018 | 32/34 | 37.2±11.8/<br>35.7±11.9 | 71.9/73.5 | MDD | DSM-IV | NA | 9.5 |
|  | Benedetti et al., | 38/19 | 46±11.1/ | 71/57.8 | BD | DSM-IV | NA | 9 |
|  | 2011 |  | 44.5±10.5 |  |  |  |  |  |
|  | Canal-Rivero et al., 2020 | 23/122 | 28.4±7.7/<br>29.4±8.3 | 39.1/37.7 | FEP | DSM-IV | NA | 9.5 |
|  | Wagner et al., 2011 | 10/15 | NR | NR | MDD | DSM-IV | NA | 8 |
| CT | Besteher et al., 2016 | 14/23 | 34.4±12.1/<br>28.8±9.7 | 57/56 | SCZ | DSM-IV | NA | 8.5 |
|  | Wagner et al., 2012 | 15/15 | 41±12.5/<br>34.1±10.5 | 41/34.1 | MDD | DSM-IV | NA | 8.5 |
|  | Taylor et al., 2015 | 21/53 | 33.5±9.1/<br>37.5±8.9 | 33.5/37.5 | MDD | DSM-IV | NA | 9 |
| rs-fMRI | Cao et al., 2016 | 35/18 | 20.6±3.6/<br>21.4±3 | 71.4/55 | MDD | DSM-IV | 1-6 months | 9.5 |
|  | Tian et al., 2021 | 42/57 | 27.3±8.3/<br>29.3±9.3 | 66.6/56.1 | BD-II | DSM-IV-TR | During episode | 9 |
|  | Cao et al., 2015 | 19/20 | 19.8±1.6/<br>20.3±1.7 | 52.6/60 | None | DSM-IV | During episode | 9.5 |
|  | Fan et al., 2013 | 27/9 | 34.4±12.9/<br>38.4±12.5 | 59.2/55.5 | MDD | DSM-IV | NA | 9.5 |
|  | Wagner et al., 2021 | 53/40 | 38±11.1/<br>37±12.2 | 85/77.5 | MDD | DSM-IV | NA | 9.5 |
|  | Gong et al., 2020 | 20/35 | 27.3±8.3/<br>23.7±7 | 65/51.4 | BD-II | DSM-IV | NA | 9 |
| FC | Kang et al., 2017 | 19/19 | 42±10.8/<br>41.1±15.2 | 57.9/47.4 | MDD | DSM-IV | NA | 9.5 |
|  | Zhang R et al., 2021 | 88/113 | 26.1±9.5/<br>28.2±9.4 | 78.4/71.7 | BD or MDD | DSM-IV | NA | 8.5 |
|  | Cheng X et al., 2020 | 30/82 | 23.5±5.3/<br>24.3±8.8 | 63.3/63.4 | BD-I | DSM-IV | NA | 8.5 |
|  | Chen Z et al., 2021 | 15/35 | 34±14/<br>35±13 | 66.6/42.8 | MDD | DSM-IV | 1 month | 8.5 |

We performed an exploratory coordinate-based meta-analysis (CBMA) using the activation likelihood estimation program GingerALE (version 3.0.2) (Eickhoff et al., 2009, pag.). The program, when supplied with the coordinates identified by the single studies, identifies commonly activated regions (clusters) of the brain across groups. CBMAs were performed for both voxel-based morphometry and resting-state index experiments that compared SA and NSA. The analysis was conducted using a cluster-level inference p < 0.05, 1000 thresholding permutations, and a cluster-forming threshold p < 0.001. The more dilated (i.e., less conservative) masking option was used. To evince any clusters that did not reach statistical significance after a cluster-level family-wise error (FWE) correction, we also conducted exploratory CBMAs without correcting for multiple comparisons (no FWE) with a cluster-forming threshold of p < 0.0001. Images were produced with BrainNet Viewer (Xia et al., 2013). Furthermore, we used JuSpace (Dukart et al., 2021) to correlate receptor spatial localization maps based on PET scans with the brain clusters identified in this meta-analysis. In this case, we implemented the default settings of the tool and compared one brain cluster with fourteen receptor maps by setting statistical significance for correlations at p < 0.0035 (Bonferroni correction).

### Functional connectivity analysis

To test the seed-based functional connectivity at rest of the areas that showed a statistically significant difference in the meta-analysis of resting-state fMRI studies, we used data from the database Human Connectome Project Young Adult (available online at https://www.humanconnectome.org/study/hcp-young-adult (Feinberg et al., 2010; Moeller et al., 2010; Setsompop et al., 2012; Van Essen et al., 2012; Xu et al., 2012)). Briefly, we selected 151 subjects (age range 22-36, 62 males) for which resting-state fMRI images were acquired with a 7 Tesla scan. Rs-fMRI data were acquired in four runs (frames per run = 900) of approximately 16 minutes each, each at the beginning of each of the four 7T imaging sessions, with eyes open with relaxed fixation on a projected bright cross-hair on a dark background (and presented in a darkened room). For the purpose of our analysis, we used data from a single run of 600 frames. Imaging parameters are: sequence = Gradient-echo EPI, TR = 1000 ms, TE = 22.2 ms, flip angle = 45 deg, FOV = 208 × 208 mm, matrix = 130 × 130, slice thickness = 1.6 mm; 85 slices; 1.6 mm isotropic voxels, multiband factor = 5, echo spacing = 0.64 ms.

RestingCstate fMRI data were preprocessed using the Connectivity Toolbox (CONN) (Whitfield-Gabrieli & Nieto-Castanon, 2012) running in Matlab R2022a (The Mathworks Inc, USA).

Functional images were realigned, slice□time corrected, spatially normalized to stereotactic space of the Montreal Neurological Institute (MNI) with a voxel size of 2×2×2□mm and spatially smoothed with a 4□mm full□width half□maximum Gaussian kernel. The Artifact Removal Tools (ART) were used to remove outlier volumes with excessive motion using liberal settings, excluding 1% of the data. Structural images were segmented into grey matter, white matter and cerebrospinal fluid and spatially normalized to MNI space. Preprocessed restin□state fMRI data were then analyzed using the CONN toolbox. Blood-oxygen-level-dependent (BOLD) data were bandpass filtered (0.008–0.09 Hz) to denoise the signal. The spatial coordinates that resulted from the meta-analysis (Talairach space – x; y; z = 53; -20; 4) were used to create a 10□mm□diameter seed.

Individual correlation maps were created by extracting the mean BOLD timecourse from the seed and calculating correlation coefficients with BOLD timecourse of each brain voxel. The value of each voxel represents the relative degree of resting-state FC with the seed region (Whitfield-Gabrieli et al., 2011). The resulting correlation map was then used for second□level analyses of FC using a one sample t-test to investigate seed□to□voxel connectivity in the group of subjects.

Moreover, we explored the correlations between the connectivity maps and scores of the relevant scales of the NIH Toolbox (life satisfaction, loneliness, friendship, purpose of life) (Hodes et al., 2013). We performed a voxel□wise statistical analysis over the entire brain using an uncorrected level (p < 1×10^−6^) and then family-wise error (FWE) correction was applied at the cluster level (p < 0.05). Permutation test (1000 simulations) was applied to test if the correlation coefficient obtained by the regression model was significantly higher than that of a random guess.

## Results

### Selected studies

A total of 5659 studies were identified from Scopus, Pubmed, and Web of Science. After duplicate removal, 3451 abstracts were screened at the title and abstract level, and 229 full-texts were retrieved for in-depth assessment. We thus excluded a total of 147 reports after full-text assessment (Figure 1), based on methodological reasons or target population (reasons for exclusion were reported in the Supplementary Appendix). Overall, we extracted data for 98 reports that met the inclusion criteria (see Methods section for further details). Some studies reported using multiple imaging modalities (e.g., VBM and rs-fMRI), and each modality accounts for a single experiment. Thus, the number of experiments considered in this systematic review and coordinate-based meta-analysis is higher (*k*=102) than the number of reports analysed. Six experiments focused on altered cortical thickness (CT), while 16 experiments used voxel-based morphometry to evince cortical, subcortical, or cerebellar structural differences. Seventy-eight experiments investigated functional differences in the target populations: twelve experiments used rs-fMRI index, 29 used fMRI while participants were performing a task, and 39 experiments investigated FC differences. Due to methodological reasons of the retrieved studies, not all experiments were considered in an exploratory CBMA. The reasons for exclusion are reported in the appropriate experiment section below. We extracted and meta-analyzed data from 22 experiments investigating the structural differences between SA and NSA. The quality of the included studies, according to an adapted version of the Imaging Methodology Quality Assessment Checklist (Supplementary Appendix and (Cattarinussi et al., 2019; Shepherd et al., 2012)), is reported in Table 1.

**Figure 1.**
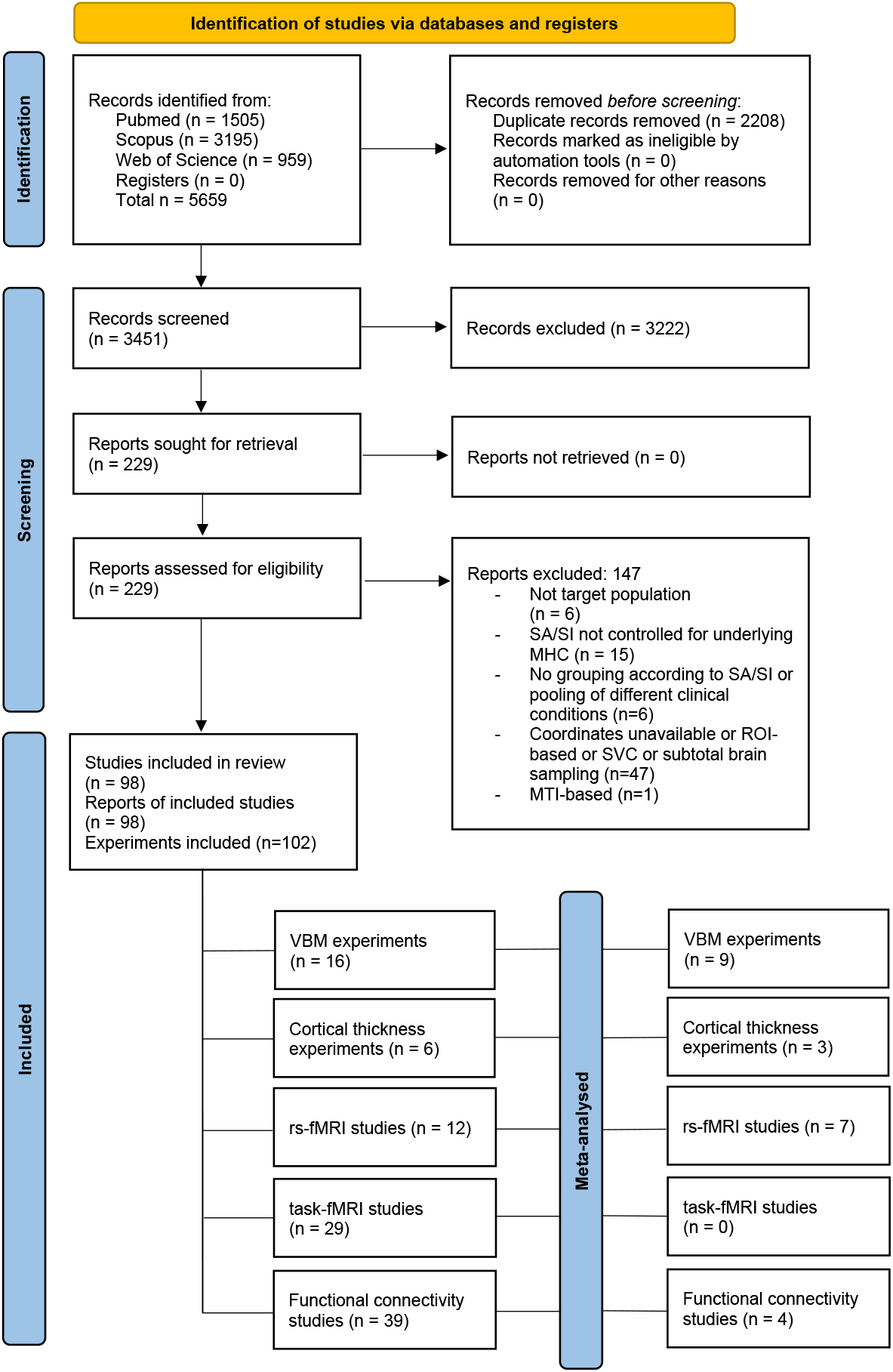
PRISMA flowchart. SA = Suicide Attempt; SI = Suicidal Ideation; MHC = Mental Health Condition; ROI = Region-of-Interest; SVC = Small Volume Correction; MTI = Magnetization Transfer Imaging; VBM = Voxel-based Morphometry; rs-fMRI = resting-state functional Mangetic Resonance Imaging.

### All-effects (Functional imaging + Morphometry) coordinate-based meta-analysis

First of all, we pooled all the resting-state fMRI, voxel-based morphometry and cortical thickness studies. Then, we meta-analysed them to identify any brain cluster of neural abnormalities across the neuroimaging modalities. We excluded the task-based studies due to the heterogeneity of tasks being adopted by the single reports, and the FC investigations for methodological reasons (eligible connectivity indexes were seed-based and thus incomparable to whole-brain indexes). A total of 18 experiments were pooled and 93 brain foci meta-analysed. This analysis comprised 1034 participants, 497 with a history of previous SA(s) and 537 NSA. However, no brain clusters were significantly different between SA and NSA in terms of neural activity at the multimodal level.

Significant brain differences between SA and NSA, according to the single imaging modality, are summarised in the following section. Table 2 recapitulates the non-statistically significant results from the CBMAs for each brain imaging modality, which are also extensively reported and discussed in the Supplementary Appendix.

**Table 2.** Summary of single imaging modality CBMAs findings. CBMA = coordinate-based meta-analysis; CT = Cortical Thickness; FC = functional connectivity; FEW = Family-wise error; GMV = grey matter volume; NSA = Non-suicide attempt; rs-fMRI = resting-state functional magnetic resonance imaging; SA = Suicide attempts; VBM = Voxel-based morphometry.

| Imaging modality | No. studies eligible | No. studies included in CBMA | Sample size (SA/NSA) | Statistical Significance | Other comments |
| --- | --- | --- | --- | --- | --- |
| VBM | 16 | 9 | 251/421 | N | All the significant volumetric differences except for two foci, identified smaller regional volumes in SA. Seven eligible studies did not find any significant |
|  |  |  |  |  | GMV differences between SA and NSA. |
| CT | 6 | 3 | 40/91 | N | Two studies did not find significant differences between the two groups after correction for multiple comparisons.<br>All the clusters (n= 7) identified by these three eligible studies reported reduced CT in SA. |
| Task-based fMRI | 29 | 0 | NA | NA | None of the task-based fMRI studies retrieved could be compared, as per methodology, to each other. |
| rs-fMRI | 11 | 6 | 196/179 | Y | A cluster belonging to the right superior temporal gyrus survived FWE correction |
| FC | 39 | 2+2 | 45/117 | N | Two studies used amygdalae as seed regions (but one found significant connectivity differences related to the left amygdala, the other one regarding the right amygdala. Thus, they could not be meta-analysed together) |

### Voxel-based morphometry findings

We extracted data for the meta-analysis from nine eligible studies, all published between 2008 and 2020, that used voxel-based morphometry. No significant spatial overlap between the reported brain regions could be identified at a p < 0.0001 with a cluster-volume of 200 mm^3^ without any statistical correction. Noteworthily, seven eligible studies (Cao et al., 2016; Duarte et al., 2017; S. Fan et al., 2019; Jia et al., 2010; Kim et al., 2015; Lippard et al., 2019; Rüsch et al., 2008) did not find any significant GMV differences between SA and NSA.

### Cortical thickness findings

Three studies that analysed CT differences between people with and without SA were eligible to be included in an exploratory CBMA. As for the CBMA based on the brain morphometry, also in this case, no significant spatial overlap between the coordinates of the CT differences could be identified at a p < 0.0001 with a cluster-volume of 200 mm^3^ (and without applying any statistical correction for multiple comparisons). Moreover, all the clusters (n= 7) identified by these three studies reported reduced CT in SA.

### Task-based fMRI findings

None of the task-based fMRI studies (*k*=29) retrieved could be compared, as per methodology, to each other, not even when pooling studies that investigated the same cognitive domain with different tasks (i.e., ignoring if the tasks employed were different – see Supplementary Appendix for reasons for exclusion of each study).

### Resting-state fMRI findings

A total of eleven studies were assessed for possible pooling in the meta-analysis. We excluded five studies due to the target population of the investigations (See Supplementary Appendix for full details). Five of the remaining six studies were conducted in China (Cao et al., 2015, 2016; T. Fan et al., 2013; Gong et al., 2020; Tian et al., 2021), only one (Wagner et al., 2021) was a multi-centre study conducted across Western countries (Canada, USA, Germany). Three studies (Cao et al., 2016; T. Fan et al., 2013; Wagner et al., 2021) included people with MDD, two studies (Gong et al., 2020; Tian et al., 2021) individuals with BD type II, and one (Cao et al., 2015) people with a history of SAs and no recognizable mental health condition. A total of 196 individuals with a history of SA(s) and of 179 diagnosis-matched patient controls were included in this exploratory meta-analysis, with a mean age ranging from 19.8 to 38.4 years old and a percentage of females in the samples ranging from 51.4 to 85%. Five studies used the ALFF as a measure of spontaneous brain activity, while one study employed regional homogeneity (ReHo) to explore local FC. A total of 37 foci coordinates were retrieved and meta-analysed: 15 foci related to spontaneous hyperactive regions in individuals with a history of suicidal behaviour, and 22 related to regions of spontaneous hypoactivation. A cluster of 520mm^3^ belonging to the right superior temporal gyrus (rSTG) (Brodmann Area 22; Talairach space coordinates – x; y; z = 53; -20; 4; Figure 2) survived FWE correction and was thus considered to be statistically significant. No brain clusters of regional hypoactivation in SA could be identified. We implemented JuSpace (Dukart et al., 2021) to correlate the coordinates of the aforementioned brain cluster with receptor spatial localisation maps based on PET scans. We found that the cluster in the rSTG was significantly correlated (R = 0.35, p = 0.003; Figure 2) with the 5-HT_1A_ receptor localisation evidenced in (Savli et al., 2012).

**Figure 2.**
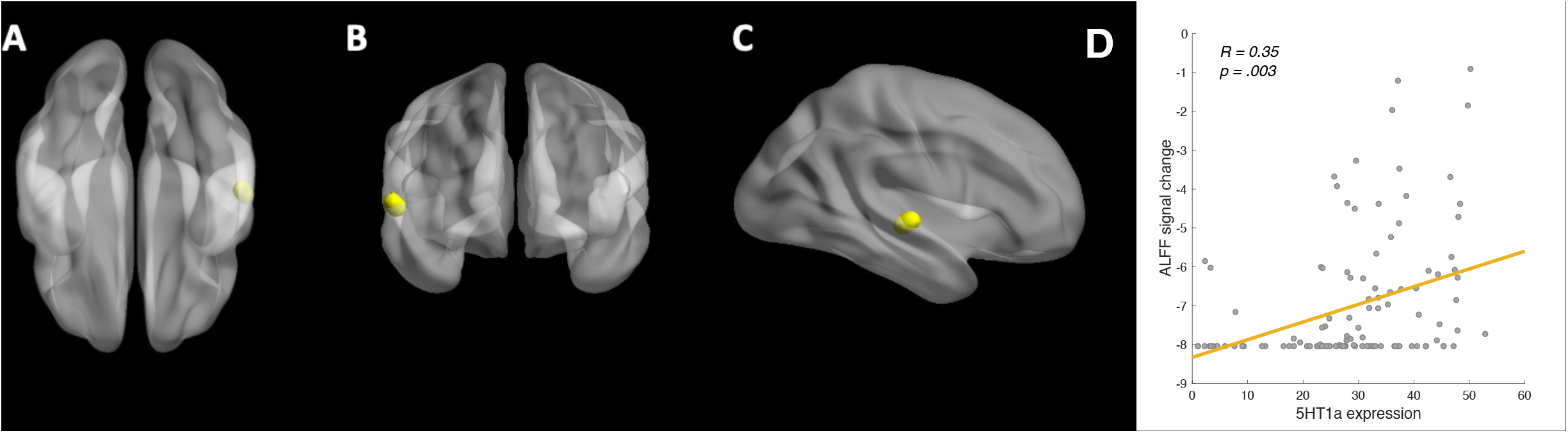
Hyperactive cluster belonging to the right superior temporal gyrus in people who attempted suicide. ALFF = Amplitude of Low-Frequency Fluctuations; 5-HT_1A_ = serotonergic (hetero)receptor 1A. A) Bottom view; B) Frontal view; C) Lateral (right) view. The signal peak belongs to Brodmann Area 22; stereotaxic Talairach coordinates – x; y; z – of the peak = [53;-20;4]) D) Correlation between 5-HT_1A_ and ALFF signal change localisation.

### Resting-state and task-based FC findings

Of the studies (*k*=39) investigating whole-brain FC that could have been considered in a CBMA, only two pairs of studies could be compared, as per methodology, to each other (see Supplementary Appendix for reasons for exclusion). However, no significant spatial overlap between the coordinates of the clusters identified from the previously cited studies could be identified at a p < 0.0001 with a cluster volume of 200 mm3 (and without applying any statistical correction for multiple comparisons).

Five studies used the bilateral amygdalae as seed regions. However, none could be compared to one another in an exploratory CBMA due to methodological reasons (see Supplementary Appendix).

### Seed-based connectivity

By leveraging the dataset from the Human Connectome Project Young Adult, we performed seed-based connectivity analyses to study the functional connections of the right STG. We observed an increase in the FC between the right STG and a bilateral set of regions spanning across fronto-insulo-temporal-occipital regions (see Supplementary Appendix for details, all p-FWE’s < 0.001). A decrease in the FC was instead observed between the right STG and the bilateral cerebellum (p-FWE < 1*10^−7^). The mean BOLD signal of the right STG presented a positive correlation with loneliness scores as measured by the NIH toolbox scales (R = 0.193, p = .018; Figure 3) and a negative correlation at trend level with friendship and purpose in life scores (R = -0.154, p = 0.059 and R = -0.147, p = 0.071, respectively). In addition, the FC of the right STG with left central operculum (MNI coordinates [x;y;z] = [-66,-1616], p-FWE = 0.02) displayed a positive correlation with the loneliness scores. Further corollary results are reported in the Supplementary Appendix and also discussed below.

**Figure 3.**
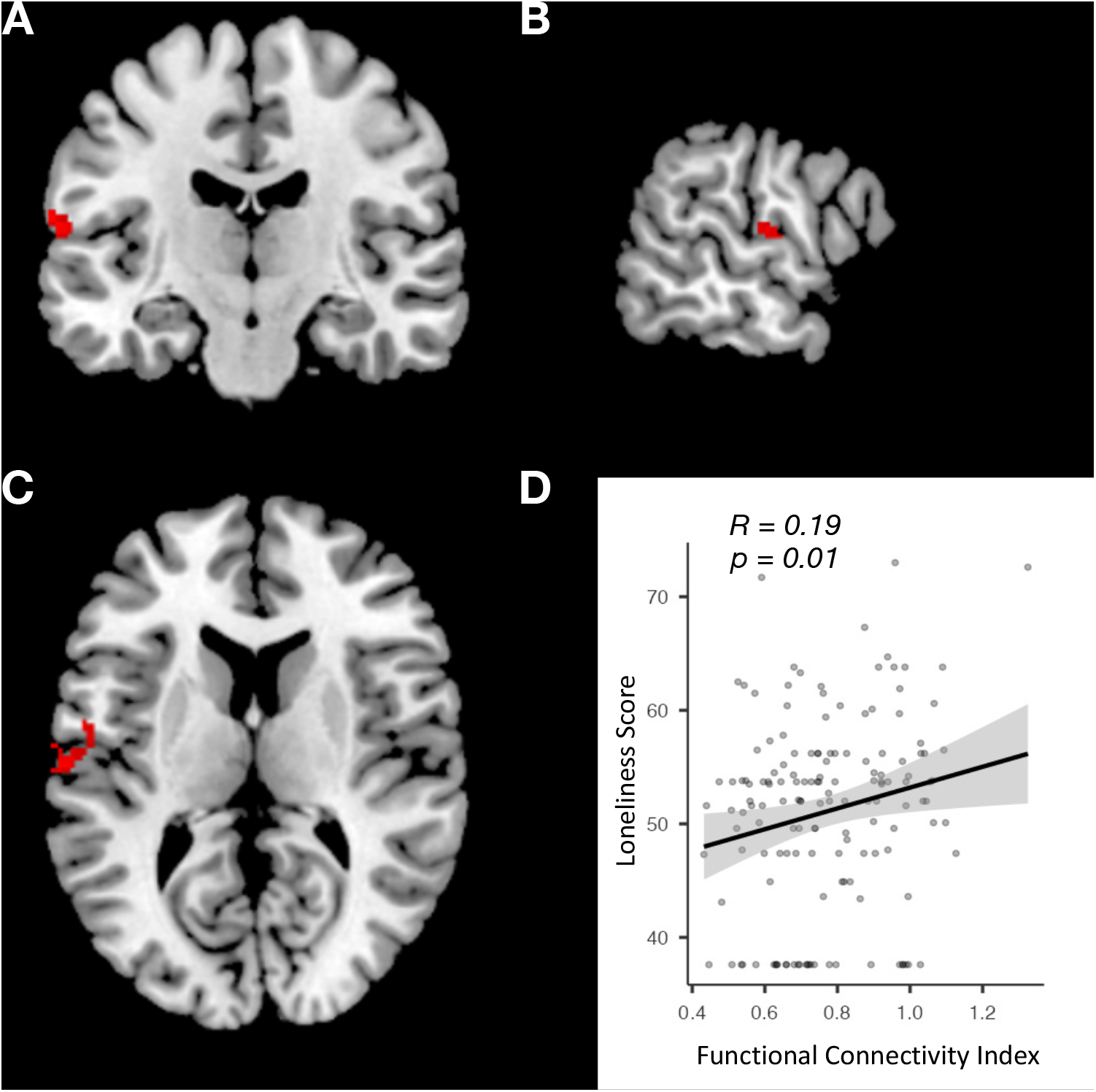
Left central operculum – right superior temporal gyrus functional connectivity correlates with loneliness scores. A) Frontal view; B) Axial view; C) Lateral view; D) Correlation plot between left central operculum to right superior temporal gyrus functional connectivity and loneliness scores as measured with NIH Toolbox.

## Discussion

SAs are preventable behaviours that usually emerge when individuals think of death as the only option to escape unbearable suffering (Pompili et al., 2008). Although several public health interventions can be put in place to prevent suicide at the population level (Hofstra et al., 2020; Hou et al., 2022; Mann et al., 2021), and psychotherapeutic (Brown et al., 2005; Büscher et al., 2020; Stanley & Brown, 2012) or pharmacological approaches (Baldessarini et al., 2006a; Zalsman et al., 2016) can be successfully implemented at the individual level, clinical tools to predict who is more likely to attempt suicide are somewhat limited, and mainly subject-dependent (Haney et al., 2012). In this context, neuroimaging markers bear the essential properties of representing fruitful objective measures of one’s risk of death by suicide (Sudol & Mann, 2017). Thus, this systematic review and exploratory CBMA aimed at investigating brain markers potentially associated with suicide attempts. We assessed for eligibility approximately 180 studies and reviewed 100 experiments regarding differences in brain morphology or function between individuals who attempted suicide and people who did not. Despite a large number of eligible studies, most results could not be compared due to methodological reasons, especially studies regarding FC and task-based fMRI. Almost all of the literature retrieved for this meta-analysis adopted a case-control study design: neuroimaging indexes of individuals who attempted suicide (SA) in the past were compared to patient controls (i.e., people that shared the same mental health condition with individuals with SA but never attempted suicide) or healthy controls ((Cao et al., 2015) in the case that no mental health condition could be identified in people with SA). The low likelihood of retrieving neuroimaging studies that prospectively investigated the neuroimaging markers of SAs is swiftly explained: SAs are rare events; this extreme behaviour is frequently the cause of first psychiatric assessment; neuroimaging studies need to have sufficient power to yield clinically meaningful results. All these factors implicate that the sample sizes required for predicting the first suicidal behaviour based on neuroimaging markers are prohibitive (larger sample sizes need to be scanned to include people who will attempt suicide).

Nonetheless, knowing if SA in the past differs from controls in neuroimaging features could inform prediction strategies. Moreover, localising such regions of difference could provide fruitful entry points to understanding the neurophysiological pathways to suicide. Lastly, a history of previous SAs is the single strongest predictor of future SAs (Berman, 2018; Christiansen & Frank Jensen, 2007; Nordström et al., 1995). This epidemiological evidence supports the view that the identification of neuroimaging markers in individuals who have already attempted suicide might become clinically helpful for objectively assessing the risk of further attempts.

*The right superior temporal gyrus as a region of interest in understanding suicidal behaviour* Our exploratory CBMA, based on 23 experiments, did not highlight any brain morphological differences among the groups of interest. However, we identified one cluster of regional hyperactivity during resting-state fMRI in the right superior temporal gyrus (rSTG) in SA. This brain region is implicated in emotion perception (Chen et al., 2021; Robins et al., 2009), spatial processing (Gharabaghi et al., 2006; Shah-Basak et al., 2018), and prediction of goal-directed movements of objects (Schultz et al., 2004) in healthy subjects, and its activity can be modulated by the antidepressant bupropion (Hama et al., 2021). In line with the evidence pinpointing this region as a hub for goal prediction, a magnetoencephalography study highlighted the possible role of the rSTG in sustaining working memory activity (Park et al., 2011). On the other hand, studies involving subjects with mental health conditions evidenced that the rSTG might play a role in illness insight (J. Fan et al., 2017) or could be a key region functionally connected to the default mode network, mediating metabolic dysfunction and mood across the lifespan (Portugal-Nunes et al., 2021). When considering those individuals who acted on their thoughts of suicide, there were also reports of morphometric differences in the rSTG (McLellan et al., 2018; Pan et al., 2015; Sarkinaite et al., 2021), whose volume can be influenced by lithium treatment (Benedetti et al., 2011; Eugene et al., 2014). Some studies eligible for this systematic review also identified volumetric reductions in the rSTG in SA compared to NSA (Benedetti et al., 2011; Canal-Rivero et al., 2020, pag.; Peng et al., 2014). However, the foci were too spatially segregated to be meta-analytically considered as belonging to the same cluster.

Similarly, two of the three eligible studies investigating CT (Besteher et al., 2016; Taylor et al., 2015) pinpointed the rSTG as a region with a statistically significant reduction of CT in SA. When considering the survivors of suicide loss, Jollant and colleagues (Jollant et al., 2018, pag. 201) highlighted a reduced volume of the temporal gyri in people with a family history of suicide, possibly indicating a neural substrate of susceptibility to transitioning from thoughts to suicidal behaviour (R. O’Connor, 2021). Regarding functional indexes of brain activation, there was another report that identified a cluster of hyperactivation in the STG of patients with a history of SAs (T. Fan et al., 2013). Yet, this regional activation was spatially distant from the two foci (Cao et al., 2016; Gong et al., 2020) composing the cluster identified by this exploratory CBMA. Moreover, there were reports of altered activation of the rSTG in individuals with non-suicidal self-injury – NSSI (Q. Huang et al., 2021; Osuch et al., 2014) and altered connectivity of this region with other brain regions during conflict and prediction errors (Harms et al., 2019; Minzenberg et al., 2015).

Pertaining the involvement of the rSTG in non-suicidal self-injury, we believe that the role of this region in the acquisition of capability to attempt suicide (i.e., transitioning from ideation – or NSSI – to SAs, as theorised in the Interpersonal Theory of Suicide (Joiner & Jr, 2005)) is a hypothesis worth of further investigation. Joiner hypothesised that the transition from suicidal ideation to a suicide attempt might also occur through physical pain insensitivity or enhanced endurance to pain (Joiner & Jr, 2005). This condition is more likely to happen as chronic self-injurious behaviours become less capable of regulating negative emotions (Grandclerc et al., 2016).

### The rSTG is enriched in 5-HT_1A_ receptors

We used JuSpace to evince the most represented receptors in the brain cluster belonging to the rSTG. We highlighted a significant spatial overlap between the localisation of 5-HT_1A_ receptors and the cluster we identified. 5-HT_1A_ can either be auto-or hetero-receptors. Autoreceptor activation plays a pivotal role in limiting the release of serotonin via a negative-feedback mechanism (Garcia-Garcia et al., 2014). In contrast, the function of heteroreceptors is less clear, although they are widely expressed in key brain regions for emotion regulation (Riad et al., 2000). It had previously been reported that people with depression who died by suicide had lower expression of 5-HT_1A_ autoreceptors in the midbrain (Boldrini et al., 2008) and were more likely to yield a polymorphism in the promoter region of the receptor gene (Lemonde et al., 2003). In the case of the localisation of this brain cluster, the spatial correlation we evidenced is more likely due to the expression of 5-HT_1A_ heteroreceptors (typically expressed in the neocortex) in the rSTG rather than autoreceptors (which are usually expressed in the midbrain – dorsal raphe nuclei). Taking these pieces of evidence together, we hypothesise that it is more likely that the putative role of the 5-HT_1A_ in this region might be dissociated from depression or anxiety severity (we would consider this alternative hypothesis if the implicated region was localised in the midbrain), but rather be associated with SA-specific dimensions, such as emotion regulation. However, to our knowledge, the role of 5-HT_1A_ heteroreceptors in suicide has not been investigated yet. Their role might have been overshadowed by the association of 5-HT_1A_ autoreceptors with depression and thus by the misconception that every person who died by suicide might have been suffering from MDD (R. O’Connor, 2021).

Given the above-mentioned and further evidence of the role of serotonergic receptors in suicide and affective disorders (i), the fact that we evidenced a spatial clustering of 5-HT_1A_ receptors in the rSTG (ii), a brain region with a role in emotion regulation, goal-directed activity (iii), and the volume of which is associated with lithium therapy (an element with potential anti-suicidal properties (Baldessarini et al., 2006b)) (iv), these four instances point at a higher likelihood that this meta-analytical finding might not be incidental. Thus, the study of 5-HT_1A_ heteroreceptors role, specifically of those expressed in the rSTG, in people who attempted suicide should be prioritised as it might inform therapeutic strategies for secondary, and hopefully primary, prevention of suicidal behaviours.

### Functional connectivity of the rSTG

The seed-based FC analysis that we conducted showed an increased FC between the right STG and several cortical and subcortical areas, suggesting that, in healthy subjects, the right STG is connected to brain regions involved in a variety of functions (see also Supplementary Appendix), spanning from visual processing (i.e. lingual gyrus, lateral occipital cortex (Clarke & Miklossy, 1990)) to motor functions (i.e. supplementary motor area (Sheets et al., 2021)), somatosensory integration (i.e. parietal operculum (Eickhoff et al., 2006)), cognitive abilities (DLPFC (Murty et al., 2011), precentral, postcentral gyri, central operculum, hippocampus (Eichenbaum, 2000; Humphreys & Lambon Ralph, 2015; Padmala & Pessoa, 2010)) and emotion regulation (i.e. insula, hippocampus (Roxo et al., 2011)). In addition, we found that resting-state activity of the rSTG presented a positive correlation with loneliness scores of the NIH toolbox and a negative correlation at the trend level with friendship and purpose in life scores. Notably, the STG is involved in several social processes, including regulation of emotional arousal, empathy and simulating the states of mind of others (i.e., theory of mind (Tholen et al., 2020)). In line with our results, altered activation of the STG has been reported in recent fMRI studies exploring neural correlates of social exclusion in association with feelings of aloneness and abandonment in individuals with borderline personality disorder (Bernheim et al., 2022). Moreover, we also observed a positive correlation between the correlation map of the right STG with the left central operculum and loneliness scores. This finding is in line with recent evidence regarding the neural correlates of the construct of perceived isolation (Spreng et al., 2020). The superior temporal regions (in particular the posterior part of the superior temporal sulcus), belonging to the default mode network (which has been linked to the subjective feeling of connection between self and others (Courtney & Meyer, 2020; Kanai et al., 2012)), and the central operculum, a hub of the cingulo-opercular network (also named the salience network), are structurally and functionally inter-related and associated with the feeling of thwarted belongingness (Layden et al., 2017). This state of loneliness can itself alter the previous brain area functions related to impulse control and vigilance against social threats (Layden et al., 2017).

Also, the correlation map of the right STG with the DLPFC displayed a positive correlation with friendship scores. Interestingly, besides its well-known role in executive functions, the DLPFC seems to be also implicated in empathic happiness (Taiwo et al., 2021) and prosocial behaviours, (Bellucci et al., 2020), so we can hypothesise that the co-activation of the right STG and the DLPFC might result in positive social functioning of the subjects and the ability to create meaningful bonds. Oppositely, a negative correlation between friendship scores and the correlation map of the right STG with the right precentral gyrus, SMA and postcentral gyrus was observed, suggesting a link between poor mentalising strategies lessened prosocial behaviours and the functional connectivity of this area (Schreuders et al., 2018).

Lastly, we found a positive correlation between the correlation map of the right STG with the right precuneus and the right DLPFC and self-efficacy scores. Interestingly, the effective rs-FC between the precuneus and the DLPFC has been proposed as a mechanism underlying social cognition (Kumar et al., 2020), a social ability in which also the STG plays a key role (Patriquin et al., 2016). In addition, an increase in rs-FC between the precuneus and the DLPFC has been proposed as the neural mechanism underlying rumination and low self-esteem in depressed patients (Cheng et al., 2018). Taken together, this evidence suggests that the rs-FC between right STG, precuneus and DLPC might be involved in social cognitive processes, and their co-activation could be associated with the development of beliefs of success in specific situations.

### Convergence of findings in light of a leading theory of suicide

As the reader might have acknowledged, we tried to contextualise the evidence that emerged from the systematic review, CBMA and functional connectivity study in light of one of the leading theories of suicide, namely Joiner’s interpersonal theory of suicide. Given the bulk of the information presented throughout the text, let us briefly recall here the main points of convergence that we found supporting the theory mentioned above. Pertinent to the results presented herein, this theory posits that the desire for suicide emerges through **thwarted belongingness** and/or perceived burdensomeness. These two prevailing ideas are commonly experienced during **depressive mental states**. The right superior temporal gyrus (rSTG), a region that we identified as possibly functionally altered in people who attempted suicide, is (i) implicated in **emotion perception** and (ii) functionally connected to the **default mode network**, which is deemed to mediate metabolic dysfunction and **mood** across the lifespan. (iii) Its volume is influenced by **lithium** treatment (the medication with the highest number of observational studies evidencing a possible anti-suicidal property), (iv). At the same time, its activity is modulated by the antidepressant **bupropion**. Coherently with the role of depression in increasing the risk of suicide, and the serotonergic pathophysiological theory for this mental health condition, we found that (v) the rSTG is significantly enriched in **5-HT**_**1A**_ **receptors**, thus further decreasing the likelihood of the statistical significance of this region being an incidental finding. With the above-reported evidence in mind, it stands out the putative role of this region in depression. Moreover, (vi) the **resting-state activity of the rSTG correlates with loneliness**, a construct strictly related to thwarted belongingness (thus showing the possible relevance of this brain area in the transition from suicidal ideation to attempt). A seed-based (rSTG-based) functional connectivity analysis evidenced that this region is functionally connected with several areas, particularly those implicated in (vii) **emotion regulation, somatosensory processing and executive functioning**. We also observed (viii) a positive correlation between the strength of the **rSTG-left central operculum connectivity and loneliness scores**, in line with the previous associations between these two areas and perceived social connection. Further supporting this finding, (ix) the strength of the **functional connection between the rSTG and DLPFC** – a region implicated in empathic happiness and social cognition – displayed a positive correlation with **self-efficacy** and **friendship** scores (a construct symmetrical to social isolation). An additional control to the latter finding is (x) the observed negative correlation between friendship scores and the connectivity of the rSTG with the motor control regions, suggesting a link to **poor mentalising** and lessened prosocial behaviour, features that undermine efficacious coping strategies.

## Limitations

Several factors limit the results of this study. Despite using a pre-defined protocol that led to the inclusion of methodologically sound studies, there were multiple sources of heterogeneity: some studies compared brain function or morphometry of individuals who attempted suicide with individuals who did not, irrespective of current suicidal ideation, whereas other studies compared people who attempted suicide and have current suicidal ideation with people with only suicidal ideation but who did not act on their thoughts. The scanners employed to acquire the brain images differed across the locations (in the manufacturer, the number of head coils, and magnetic field strength), and so did the pipelines of analysis that the authors applied. Moreover, the time from suicide attempts to scan varied across the studies (although most articles did not report this information): this could be why some studies did not identify differences in brain functionality of people who attempted suicide in their lifetime with respect to patient controls. Regarding the heterogeneity in the demographics of the samples being studied, mean age varied greatly (and accordingly to the population of reference, i.e., younger patients with first-episode psychoses and older patients with major depressive disorders were included in this meta-analysis), as well as the criteria of inclusion/exclusion of individuals with active or past alcohol/substance use disorders (AUD/SUD, most of the studies enrolled individuals with a history of AUD/SUD as long as the subjects had been unexposed to substances for the last 1-6 months before scan). Lastly, a consideration on the survival bias needs to be mentioned: half of the individuals who die by suicide lose their life at their first attempt. Thus, it is impossible to know if the population that loses their lives at the first attempt differs in brain functionality from those who survive.

## Conclusions

Taken together, this meta-analysis pinpoints a cluster in the rSTG, enriched with 5-HT_1A_ receptors, the activity of which differs between individuals with a history of suicide attempts and those without. This cluster of the rSTG needs to be replicated by further studies, as it derives from the analysis of individuals from East Asia, for whom differences in the morphometry of the right superior temporal gyrus have been reported (D. W. Kang et al., 2020). Furthermore, we evidenced that the activity of this area and its functional connectivity strength with the left central operculum is significantly correlated with the perceived loneliness score as measured by the NIH Toolbox, thus providing a putative neural substrate mediating the sense of thwarted belongingness, which entails a stronger desire for death as postulated by Joiner’s Interpersonal Theory of Suicide. Moreover, this systematic review highlighted that the rSTG could represent a brain region with reduced grey matter volume in people who attempted suicide. However, the foci of morphological difference identified by the single studies are too spatially distant to be included in the same brain cluster. Future research should consider the presence of suicidal ideation and its severity, apply statistical corrections at the whole-brain level, employ reproducible and verified pipelines of analysis, and be based on a pre-registered protocol with a detailed power analysis.

## Data Availability

All data produced in the present work are contained in the manuscript

## Acknowledgements

We wish to thank Prof. Rory O’Connor for the publication of his book “When it is darkest – Why people die by suicide and what we can do to prevent it” (R. O’Connor, 2021). Data were provided [in part] by the Human Connectome Project, WU-Minn Consortium (Principal Investigators: David Van Essen and Kamil Ugurbil; 1U54MH091657), funded by the 16 NIH Institutes and Centers that support the NIH Blueprint for Neuroscience Research; and by the McDonnell Center for Systems Neuroscience at Washington University.

## Author contributions

Conceptualisation: NM

Methodology: NM, AM, GC, FS

Investigation: NM, AM, GC, FS

Formal analysis: NM, GC, FS

Visualisation: NM, GC

Supervision: FS

Writing – original draft: NM, AM

Writing – review & editing: NM, AM, GC, FS

## Competing interests None

## Data and materials availability

All data can be retrieved from the manuscript or supplementary appendix.

