## Supplementary Material for "Whole-brain structural and functional neuroimaging of individuals who attempted suicide and people who did not: a systematic review, exploratory coordinate-based meta-analysis and seed-based connectivity study"

**INDEX**

PRISMA Checklist …………………………………………………………………………………...3

PRISMA Abstract Checklist………………………………………………………………………….7

Supplementary Table S1……………………………………………………………………………...8

Supplementary Table S2…………………………………………………………………………….12

Supplementary Table S3…………………………………………………………………………….15

Supplementary Table S4…………………………………………………………………………….16

Supplementary Results ……………………………………………………………………………...17

Supplementary Figure S1……………………………………………………………………………23

Supplementary References ………………………………………………………………………….24

**PRISMA Checklist**

| **Topic** | **No.** | **Item** | **Location where item is reported** |
| --- | --- | --- | --- |
| **TITLE** |  |  |  |
| **Title** | 1 | Identify the report as a systematic review. | Page 1 |
| **ABSTRACT** |  |  |  |
| **Abstract** | 2 | See the PRISMA 2020 for Abstracts checklist | Suppl. Appendix |
| **INTRODUCTION** |  |  |  |
| **Rationale** | 3 | Describe the rationale for the review in the context of existing knowledge. | Page 4-6 |
| **Objectives** | 4 | Provide an explicit statement of the objective(s) or question(s) the review addresses. | Page 5-6 |
| **METHODS** |  |  |  |
| **Eligibility criteria** | 5 | Specify the inclusion and exclusion criteria for the review and how studies were grouped for the syntheses. | Page 7 |
| **Information sources** | 6 | Specify all databases, registers, websites, organisations, reference lists and other sources searched or consulted to identify studies. Specify the date when each source was last searched or consulted. | Page 6 |
| **Search strategy** | 7 | Present the full search strategies for all databases, registers and websites, including any filters and limits used. | Page 6 |
| **Selection process** | 8 | Specify the methods used to decide whether a study met the inclusion criteria of the review, including how many reviewers screened each record and each report retrieved, whether they worked independently, and if applicable, details of automation tools used in the process. | Page 6-7 |
| **Data collection process** | 9 | Specify the methods used to collect data from reports, including how many reviewers collected data from each report, whether they worked independently, any processes for obtaining or confirming data from study investigators, and if applicable, details of automation tools used in the process. | Page 8-9 |
| **Data items** | 10a | List and define all outcomes for which data were sought. Specify whether all results that were compatible with each outcome domain in each study were sought (e.g. for all measures, time points, analyses), and if not, the methods used to decide which results to collect. | Page 8 |
|  | 10b | List and define all other variables for which data were sought (e.g. participant and intervention characteristics, funding sources). Describe any assumptions made about any missing or unclear information. | Page 7 |
| **Study risk of bias assessment** | 11 | Specify the methods used to assess risk of bias in the included studies, including details of the tool(s) used, how many reviewers assessed each study and whether they worked independently, and if applicable, details of automation tools used in the process. | Page 8-9 |
| **Effect measures** | 12 | Specify for each outcome the effect measure(s) (e.g. risk ratio, mean difference) used in the synthesis or presentation of results. | Page 9-11 |
| **Synthesis methods** | 13a | Describe the processes used to decide which studies were eligible for each synthesis (e.g. tabulating the study intervention characteristics and comparing against the planned groups for each synthesis (item 5)). | Page 7-11 |
|  | 13b | Describe any methods required to prepare the data for presentation or synthesis, such as handling of missing summary statistics, or data conversions. | Page 9-11 |
|  | 13c | Describe any methods used to tabulate or visually display results of individual studies and syntheses. | Page 8-11 |
|  | 13d | Describe any methods used to synthesize results and provide a rationale for the choice(s). If meta-analysis was performed, describe the model(s), method(s) to identify the presence and extent of statistical heterogeneity, and software package(s) used. | Page 8-11 |
|  | 13e | Describe any methods used to explore possible causes of heterogeneity among study results (e.g. subgroup analysis, meta-regression). | N.A. |
|  | 13f | Describe any sensitivity analyses conducted to assess robustness of the synthesized results. | N.A. |
| **Reporting bias assessment** | 14 | Describe any methods used to assess risk of bias due to missing results in a synthesis (arising from reporting biases). | Page 9 |
| **Certainty assessment** | 15 | Describe any methods used to assess certainty (or confidence) in the body of evidence for an outcome. | N.A. |
| **RESULTS** |  |  |  |
| **Study selection** | 16a | Describe the results of the search and selection process, from the number of records identified in the search to the number of studies included in the review, ideally using a flow diagram. | Page 11 |
|  | 16b | Cite studies that might appear to meet the inclusion criteria, but which were excluded, and explain why they were excluded. | Page 11-14, Suppl. Appendix |
| **Study characteristics** | 17 | Cite each included study and present its characteristics. | Page 11-14, Table 1 and Suppl. Appendix |
| **Risk of bias in studies** | 18 | Present assessments of risk of bias for each included study. | Suppl. Appendix |
| **Results of individual studies** | 19 | For all outcomes, present, for each study: (a) summary statistics for each group (where appropriate) and (b) an effect estimate and its precision (e.g. confidence/credible interval), ideally using structured tables or plots. | Table 1, Suppl. Appendix |
| **Results of syntheses** | 20a | For each synthesis, briefly summarise the characteristics and risk of bias among contributing studies. | Suppl. Appendix |
|  | 20b | Present results of all statistical syntheses conducted. If meta-analysis was done, present for each the summary estimate and its precision (e.g. confidence/credible interval) and measures of statistical heterogeneity. If comparing groups, describe the direction of the effect. | Page 11-14 |
|  | 20c | Present results of all investigations of possible causes of heterogeneity among study results. | Page 11-14 |
|  | 20d | Present results of all sensitivity analyses conducted to assess the robustness of the synthesized results. | N.A. |
| **Reporting biases** | 21 | Present assessments of risk of bias due to missing results (arising from reporting biases) for each synthesis assessed. | N.A. |
| **Certainty of evidence** | 22 | Present assessments of certainty (or confidence) in the body of evidence for each outcome assessed. | N.A. |
| **DISCUSSION** |  |  |  |
| **Discussion** | 23a | Provide a general interpretation of the results in the context of other evidence. | Page 15-20 |
|  | 23b | Discuss any limitations of the evidence included in the review. | Page 21-22 |
|  | 23c | Discuss any limitations of the review processes used. | Page 21-22 |
|  | 23d | Discuss implications of the results for practice, policy, and future research. | Page 20-22 |
| **OTHER INFORMATION** |  |  |  |
| **Registration and protocol** | 24a | Provide registration information for the review, including register name and registration number, or state that the review was not registered. | Page 6 |
|  | 24b | Indicate where the review protocol can be accessed, or state that a protocol was not prepared. | Page 6 |
|  | 24c | Describe and explain any amendments to information provided at registration or in the protocol. | N.A. |
| **Support** | 25 | Describe sources of financial or non-financial support for the review, and the role of the funders or sponsors in the review. | Page 23 |
| **Competing interests** | 26 | Declare any competing interests of review authors. | Page 33 |
| **Availability of data, code and other materials** | 27 | Report which of the following are publicly available and where they can be found: template data collection forms; data extracted from included studies; data used for all analyses; analytic code; any other materials used in the review. | Page 33 |

**PRISMA Abstract Checklist**

| **Topic** | **No.** | **Item** | **Reported?** |
| --- | --- | --- | --- |
| **TITLE** |  |  |  |
| **Title** | 1 | Identify the report as a systematic review. | Yes |
| **BACKGROUND** |  |  |  |
| **Objectives** | 2 | Provide an explicit statement of the main objective(s) or question(s) the review addresses. | Yes |
| **METHODS** |  |  |  |
| **Eligibility criteria** | 3 | Specify the inclusion and exclusion criteria for the review. | Yes |
| **Information sources** | 4 | Specify the information sources (e.g. databases, registers) used to identify studies and the date when each was last searched. | Yes |
| **Risk of bias** | 5 | Specify the methods used to assess risk of bias in the included studies. | No |
| **Synthesis of results** | 6 | Specify the methods used to present and synthesize results. | No |
| **RESULTS** |  |  |  |
| **Included studies** | 7 | Give the total number of included studies and participants and summarise relevant characteristics of studies. | Yes |
| **Synthesis of results** | 8 | Present results for main outcomes, preferably indicating the number of included studies and participants for each. If meta-analysis was done, report the summary estimate and confidence/credible interval. If comparing groups, indicate the direction of the effect (i.e. which group is favoured). | Yes |
| **DISCUSSION** |  |  |  |
| **Limitations of evidence** | 9 | Provide a brief summary of the limitations of the evidence included in the review (e.g. study risk of bias, inconsistency and imprecision). | Yes |
| **Interpretation** | 10 | Provide a general interpretation of the results and important implications. | Yes |
| **OTHER** |  |  |  |
| **Funding** | 11 | Specify the primary source of funding for the review. | No |
| **Registration** | 12 | Provide the register name and registration number. | No |

| **First author, year** | **Imaging Power Field** | **Type of analysis** | **Contrast** | **Peak voxel localization (MNI)** | | |
| --- | --- | --- | --- | --- | --- | --- |
| Wang, 2020 | 3 T | VBM | NSA>SA | **x** | **y** | **z** |
|  |  |  |  | 28,5 | 39 | 37,5 |
|  |  |  |  | -25,5 | 31,5 | 42 |
| Peng, 2014 | 3 T | VBM | NSA>SA | **x** | **y** | **z** |
|  |  |  |  | -2 | -21 | 28 |
| Johnston, 2017 | 3 T | VBM | NSA>SA | **x** | **y** | **z** |
|  |  |  |  | 16 | 36 | -24 |
|  |  |  |  | 28 | -34 | 8 |
|  |  |  |  | -4 | -80 | -18 |
| Lee, 2016 | 1.5 T | VBM | NSA>SA | **x** | **y** | **z** |
|  |  |  |  | -50 | -73 | 17 |
|  |  |  |  | 14 | -68 | -34 |
| Aguilar, 2008 | 1.5 T | VBM | NSA>SA | **x** | **y** | **z** |
|  |  |  |  | -41.9 | -30.52 | 11.47 |
|  |  |  |  | -9.14 | 40.72 | -31.99 |
| Jollant, 2018 | 3 T | VBM | NSA>SA | **x** | **y** | **z** |
|  |  |  |  | 2 | 12 | 50 |
| Benedetti, 2011 | 3 T | VBM | NSA>SA | **x** | **y** | **z** |
|  |  |  |  | -12 | 55 | 33 |
|  |  |  |  | -4 | 8 | 53 |
|  |  |  |  | -8 | 65 | 20 |
|  |  |  |  | -16 | 19 | 63 |
|  |  |  |  | -30 | 19 | 54 |
|  |  |  |  | -49 | 51 | -4 |
|  |  |  |  | -33 | 59 | 8 |
|  |  |  |  | -24 | 34 | -12 |
|  |  |  |  | -11 | 35 | -28 |
|  |  |  |  | -35 | 31 | -20 |
|  |  |  |  | -9 | -30 | 56 |
|  |  |  |  | 23 | 42 | 41 |
|  |  |  |  | 4 | 17 | 59 |
|  |  |  |  | 27 | 17 | 59 |
|  |  |  |  | 6 | 50 | 41 |
|  |  |  |  | -43 | -4 | 18 |
|  |  |  |  | 70 | -22 | 7 |
|  |  |  |  | 50 | -55 | -18 |
|  |  |  |  | -4 | 44 | 7 |
|  |  |  |  | -3 | 19 | 24 |
|  |  |  |  | 4 | 43 | 10 |
|  |  |  |  | 5 | 26 | 34 |
|  |  |  |  | -38 | -60 | 51 |
|  |  |  |  | -6 | -53 | 65 |
|  |  |  |  | -52 | -66 | 37 |
|  |  |  |  | -2 | -32 | 54 |
|  |  |  |  | -2 | -19 | 50 |
|  |  |  |  | 50 | -34 | 54 |
|  |  |  |  | 55 | -61 | 40 |
|  |  |  |  | -7 | -71 | -9 |
|  |  |  |  | -52 | -72 | 0 |
|  |  |  |  | -2 | -82 | 31 |
|  |  |  |  | -3 | -44 | 45 |
|  |  |  |  | -28 | -94 | -20 |
|  |  |  |  | 41 | -78 | 31 |
|  |  |  |  | 36 | -44 | -13 |
|  |  |  |  | 8 | -30 | 8 |
|  |  |  |  | -16 | 18 | 6 |
|  |  |  | SA>NSA | **x** | **y** | **z** |
|  |  |  |  | -45 | -33 | 6 |
|  |  |  |  | 50 | -43 | 16 |
| Canal-Rivero, 2020 | 3 T | VBM | NSA>SA | **x** | **y** | **z** |
|  |  |  |  | 57 | 8 | 10 |
|  |  |  |  | 12 | 30 | 36 |
|  |  |  |  | -4 | 6 | -14 |
|  |  |  |  | -24 | -4 | 45 |
|  |  |  |  | 22 | 9 | -28 |
|  |  |  |  | -15 | -94 | 2 |
|  |  |  |  | -14 | -28 | 50 |
|  |  |  |  | -44 | -8 | 38 |
|  |  |  |  | 14 | -40 | 45 |
|  |  |  |  | 0 | 50 | 20 |
|  |  |  |  | 28 | 42 | 24 |
|  |  |  |  | -27 | -9 | 64 |
|  |  |  |  | 9 | 22 | -8 |
|  |  |  |  | 10 | -4 | -26 |
| Wagner, 2011 | 1.5 T | VBM | NSA>SA | **x** | **y** | **z** |
|  |  |  |  | 14.95 | 22.36 | -11.54 |
| Besteher, 2016 | 1.5 T | Cortical Thickness | NSA>SA | **x** | **y** | **z** |
|  |  |  |  | 44.25 | 14.33 | -37.12 |
| Wagner, 2012 | 1.5 T | Cortical Thickness | NSA>SA | **x** | **y** | **z** |
|  |  |  |  | -49.9 | 31.81 | -10.25 |
|  |  |  |  | -41 | 8 | 18 |
| Taylor, 2015 | 3 T | Cortical Thickness | NSA>SA | **x** | **y** | **z** |
|  |  |  |  | -33,5 | 1,4 | 6,5 |
|  |  |  |  | -24,5 | 4,1 | 42,9 |
|  |  |  |  | -31,3 | -46,7 | 48,2 |
|  |  |  |  | -52,6 | -10,6 | 0,4 |
| Cao, 2016 | 3 T | zALFF (rs-fMRI) | NSA>SA | **x** | **y** | **z** |
|  |  |  |  | -33 | 63 | -3 |
|  |  |  |  | -34 | 59 | -3 |
|  |  |  | SA>NSA | **x** | **y** | **z** |
|  |  |  |  | -66 | -51 | -3 |
|  |  |  |  | 63 | -15 | 3 |
|  |  |  |  | -51 | -60 | 9 |
|  |  |  |  | -51 | -66 | 27 |
| Tian, 2021 | 3 T | sALFF (rs-fMRI) | SA>NSA | **x** | **y** | **z** |
|  |  |  |  | 0 | 33 | 21 |
| Cao, 2015 | 3 T | ReHo  (rs-fMRI) | NSA>SA | **x** | **y** | **z** |
|  |  |  |  | 22 | -48 | -50 |
|  |  |  |  | 44 | -73 | -40 |
|  |  |  |  | -21 | -70 | -20 |
|  |  |  |  | -23 | -32 | -20 |
|  |  |  |  | -22 | -16 | -20 |
|  |  |  |  | -21 | -32 | -10 |
|  |  |  |  | 26 | -30 | -10 |
|  |  |  |  | -36 | 25 | -10 |
|  |  |  |  | -25 | 46 | 30 |
|  |  |  |  | 36 | 47 | 30 |
|  |  |  |  | 33 | -63 | 50 |
| Fan, 2013 | 3 T | mALFF (rs-fMRI) | NSA>SA | **x** | **y** | **z** |
|  |  |  |  | 9 | 57 | -12 |
|  |  |  | SA>NSA | **x** | **y** | **z** |
|  |  |  |  | 45 | -66 | 24 |
| Wagner, 2021 | 3 T | zALFF (rs-fMRI) | SI>SA | **x** | **y** | **z** |
|  |  |  |  | -46 | -62 | 32 |
|  |  |  |  | -48 | -32 | 36 |
|  |  |  |  | -38 | -74 | 24 |
|  |  |  |  | 50 | -54 | 34 |
|  |  |  |  | 14 | -70 | 46 |
|  |  |  |  | 56 | -44 | 38 |
|  |  |  |  | 24 | -58 | 64 |
|  |  |  |  | 18 | -44 | 72 |
|  |  |  | SA>SI | **x** | **y** | **z** |
|  |  |  |  | -36 | -16 | -30 |
|  |  |  |  | 42 | -12 | -36 |
|  |  |  |  | -22 | -22 | -10 |
| Gong, 2020 | 3 T | dALFF (rs-fMRI | SA>SI | **x** | **y** | **z** |
|  |  |  |  | 36 | 3 | -36 |
|  |  |  |  | 51 | -69 | 0 |
|  |  |  |  | 60 | -21 | 0 |
| Yang, 2022 | 3 T | ReHo | SA>NSA | **x** | **y** | **z** |
|  |  |  |  | 3 | -42 | 15 |
| Kang, 2017 | 1.5T | FC (seed: amigdala) | SA>NSA | **x** | **y** | **z** |
|  |  |  |  | -54 | -15 | -15 |
| Zhang R, 2021 | 3 T | FC (seed: amigdala) | SA>NSA | **x** | **y** | **z** |
|  |  |  |  | -9 | -42 | 72 |
|  |  |  |  | 12 | -45 | 69 |
| Cheng, 2020 | 3 T | Whole-brain FC | SA>NSA | **x** | **y** | **z** |
|  |  |  |  | -51 | -27 | 9 |
|  |  |  | NSA>SA | **x** | **y** | **z** |
|  |  |  |  | 39 | -12 | 21 |
|  |  |  |  | -39 | -18 | 9 |
|  |  |  |  | 12 | -84 | 45 |
| Chen, 2021 | 3 T | Whole-brain FC | SA>NSA | **x** | **y** | **z** |
|  |  |  |  | 42 | 45 | 0 |
|  |  |  |  | 3 | 39 | 48 |

**Supplementary Table S1. Imaging acquisition characteristics and cluster coordinates identified by the studies included in the coordinate-based meta-analysis.** ALFF = Amplitude of Low-Frequency Fluctuations; dALFF = dynamic ALFF; mALFF = global mean ALFF; sALFF = static ALFF; zALFF = normalized ALFF; FC = functional connectivity; MNI = Montreal Neurological Institute; NSA = Individuals without a history of Suicide Attempt; ReHo = Regional Homogeneity; SA = Individuals with a history of Suicide Attempt(s); SI = Individuals with Suicidal Ideation at the time of scanning; VBM = Voxel-based Morphometry;

| Type of analysis | DOI | First author, year | Reason for exclusion |
| --- | --- | --- | --- |
| VBM | 10.1017/S0954579418000822 | Beauchaine, 2019 | NSSI |
|  | 10.1017/S0954579418000822 | Fan, 2019 | No GMV differences |
|  | 10.1016/j.jad.2018.11.097 | Lippard, 2019 | ROI-based |
|  | 10.1016/j.pscychresns.2017.04.012 | Duarte, 2017 | No GMV differences |
|  | 10.1016/j.jad.2016.07.002 | Cao, 2016 | No GMV differences |
|  | 10.1038/srep09670 | Chen, 2015 | MTI-based |
|  | 10.1016/j.jad.2015.01.001 | Kim, 2015 | No GMV differences |
|  | 10.1007/s00429-013-0562-2 | Duerden, 2014 | NSSI |
|  | [10.1176/appi.ajp.2010.09101513](https://doi.org/10.1176/appi.ajp.2010.09101513) | Jia, 2012 | No GMV differences |
|  | 10.1177/0891988710363713 | Hwang, 2010 | No slice number/thickness |
|  | 10.1016/j.pscychresns.2007.12.011 | Rüsch, 2008 | No GMV differences |
| Cortical thickness | 10.1016/j.pscychresns.2020.111032 | Kang, 2020 | No thickness differences |
|  | 10.1016/j.jad.2018.10.081 | Segreti, 2019 | No thickness differences |
|  | 10.1192/bjp.bp.114.151316 | Pan, 2015 | Only adolescents |
| rs-fMRI | 10.1016/j.pnpbp.2021.110253 | Athanassiou, 2021 | Patient controls grouped with healthy controls |
|  | 10.1176/appi.ajp.2020.20020120 | Vidal-Ribas, 2021 | Only children |
|  | 10.4103/1673-5374.274346 | Huang, 2020 | Only adolescents |
|  | 10.1002/da.22888 | Lan, 2019 | Only suicidal ideation |
|  | 10.1017/S0033291718001502 | Li, 2019 | Only suicidal ideation |
| Task-fMRI | 10.1016/j.bpsc.2020.11.004 | Mayo, 2021 | NSSI |
|  | 10.1176/appi.ajp.2020.20020120 | Vidal-Ribas, 2021 | Only children |
|  | 10.1016/j.bpsc.2019.10.016 | Dir, 2020 | Only children |
|  | 10.1016/j.pscychresns.2019.05.001 | Harms, 2019 | Only adolescents |
|  | 10.1016/j.eclinm.2019.06.016 | Perini, 2019 | NSSI |
|  | 10.1016/j.jpsychires.2018.08.018 | Ai, 2018 | Only eligible study of emotion recognition |
|  | 10.1016/j.bpsc.2017.08.008 | Miller, 2018 | Only adolescents |
|  | 10.1155/2018/9898654 | Potvin, 2018 | Only eligible study with risk-taking paradigm |
|  | 10.1007/s11682-017-9687-x | Vega, 2018 | NSSI |
|  | 10.1038/s41598-017-10541-5 | Baek, 2017 | No functional index differences |
|  | 10.1016/j.pscychresns.2016.08.001 | Groschwitz, 2016 | NSSI |
|  | 10.1038/s41598-017-00211-x | Olié, 2017 | Only eligible study with social exclusion paradigm |
|  | 10.1017/S0033291715002421 | Richard-Devantoy, 2016 | No functional index differences |
|  | 10.1016/j.jpsychires.2016.06.020 | Silvers, 2016 | Cerebellum not included |
|  | 10.1017/S0033291715001890 | Vanyukov, 2016 | Only eligible study with delayed reward paradigm |
|  | 10.1016/j.pnpbp.2015.03.005 | Lee, 2015 | NSSI |
|  | 10.1017/S0954579415000449 | Sauder, 2015 | NSSI |
|  | 10.1037/a0036962 | Davis, 2014 | NSSI |
|  | 10.1016/j.schres.2014.05.039 | Minzenberg, 2014 | Cerebellum not included |
|  | 10.1016/j.pscychresns.2014.05.003 | Osuch, 2014 | NSSI |
|  | 10.1016/j.pscychresns.2012.07.008 | Pan, 2013a | Only adolescents |
|  | 10.1017/S0033291712002966 | Pan, 2013b | Only adolescents |
|  | 10.1097/PSY.0b013e31824f888f | Matthwes, 2012 | Only suicidal ideation |
|  | 10.1016/j.pscychresns.2011.12.012 | Plener, 2012 | NSSI |
|  | 10.1016/j.jaac.2011.03.018 | Pan, 2011 | Only adolescents |
|  | 10.1016/j.neuroimage.2010.03.027 | Jollant, 2010 | No functional index differences |
|  | 10.1176/appi.ajp.2008.07081239 | Jollant, 2008 | Signal related to neutral faces subtracted to signal of angry faces |
| FC | 10.1016/j.pnpbp.2021.110253 | Athanassiou, 2021 | Patient controls grouped with healthy controls |
|  | 10.1007/s10548-021-00830-8 | Hu, 2021 | Only eligible study with seed in the Insula |
|  | 10.1038/s41398-020-01103-x | Ho, 2021 | Only adolescents |
|  | 10.1016/j.bbr.2020.112544 | Qiu, 2020 | Only eligible study with seed in the ACC |
|  | 10.3389/fpsyt.2020.597770 | Zhu, 2020 | Only eligibile study with seed in amygdala and FC of Fronto-Limbic circuits |
|  | 10.3389/fpsyt.2019.00923 | Wang, 2020a | Only eligibile study with seed in amygdala and FC of Prefrontal Cortex |
|  | 10.1038/s41386-020-0632-0 | Brown, 2020 | Only eligibile study of task-based FC (decision-making) |
|  | 10.1016/j.psychres.2019.112713 | Cao, 2020 | Same population of * |
|  | 10.1017/S0033291719001132 | Malhi, 2020 | No functional index differences |
|  | 10.1016/j.jad.2020.07.016 | Wang, 2020b | No functional index differences |
|  | 10.3389/fpsyt.2020.608197 | Qiao, 2020 | Only suicidal ideation |
|  | 10.1002/jmri.27499 | Li, 2020 | Only eligibile study on morphological networks |
|  | 10.1111/acps.13029 | Gosnell, 2019 | Coordinates of seeds not provided |
|  | 10.1176/appi.neuropsych.17120351 | Ambrosi, 2019 | Only eligible study with seed in the Habenula |
|  | 10.1038/s41598-019-50881-y | Wagner, 2019 | Only eligible study of network topology |
|  | 10.3389/fpsyt.2019.00044 | Barredo, 2019 | Only suicidal ideation |
|  | 10.1111/sltb.12471 | Schreiner, 2019 | Only adolescents |
|  | 10.1016/j.jaac.2018.06.036 | Alarcón, 2019 | Only adolescents |
|  | 10.1089/cap.2018.0152 | Santamarina-Perez, 2019 | NSSI |
|  | 10.4088/JCP.17m11901 | Cáceda, 2018 | Cases grouped with controls |
|  | 10.1186/s12991-018-0208-0 | Wei, 2018 | Cases grouped with controls |
|  | 10.1016/j.jad.2017.09.021 | Ordaz, 2018 | Only suicidal ideation |
|  | 10.1002/hbm.24235 | Liao, 2018 | Only suicidal ideation |
|  | 10.1176/appi.ajp.2016.15050652 | Johnston, 2017 | Only eligibile study of task-based FC (face recognition) |
|  | 10.1038/s41598-017-15926-0 | Kim, 2017 | Cases grouped with controls |
|  | 10.1016/j.jad.2017.06.004 | Schreiner, 2017 | NSSI |
|  | 10.1016/j.jad.2017.02.027 | Du, 2017 | Only suicidal ideation |
|  | 10.1176/appi.neuropsych.15120422 | Minzenberg, 2016 | Both meta-analysed, no clusters identified |
|  | 10.1016/j.jpsychires.2015.04.002 | Minzenberg, 2015 |  |
|  | 10.1016/j.jad.2013.01.028 | Marchand, 2013 | Only eligible study with seed in the pCC |
|  | 10.1016/j.pnpbp.2011.10.016 | Marchand, 2012 | Only eligible study with seed in the Putamen |
|  | 10.1016/j.pnpbp.2017.04.029 | Kang, 2017 | Only eligible study with amygdala to whole-brain connectivity |
|  | 10.3389/fnhum.2020.585664 | Zhang R, 2021 | Pooling of different suicidal conditions |
|  | 10.1186/s12888-016-1047-7 | Zhang S, 2016 | Only eligible study employing ICA for DMN connectivity |
|  | 10.1038/s41386-019-0560-z | Jung, 2020 | Only eligible study employing ICA for intra-network connectivity |
|  | 10.1017/S0033291719002356 | Stange, 2020 | Seed localizations not overlapping with other eligible studies |
|  | 10.1111/bdi.13012 | Cheng, 2020 | Only eligible study employing global brain connectivity measures |
|  | 10.1016/j.jad.2020.11.061 | Stumps, 2021 | Only eligible study employing hub indexes of network nodes |

**Supplementary Table S2. Characteristics of the experiments excluded from the coordinate-based meta-analysis and reasons for exclusion.**

ACC = Anterior Cingulate Cortex; FC = Functional Connectivity; GMV = Grey Matter Volume; MTI = Magnetization Transfer Imaging; NSSI = Non-suicidal Self-Injury; pCC = posterior Cingulate Cortex; ROI = Region-of-Interest; rs-fMRI = resting-state functional Magnetic Resonance Imaging; VBM = Voxel-based Morphometry; Studies that did not identify differences in the neuroimaging features need to be excluded from the meta-analysis: the methodological reasons for doing so are explained in ^1,2^; * = 10.1186/s12888-016-1047-7 (Zhang S et al., 2016).

| Study | Risk of bias assessment | | | |
| --- | --- | --- | --- | --- |
|  | Subjects | Methods | Results & Conclusions | Overall |
| Wang et al., 2020 | - | + | + | - |
| Peng et al., 2014 | - | + | + | - |
| Johnston et al., 2017 | + | + | + | + |
| Lee et al., 2016 | - | + | + | - |
| Aguilar et al., 2008 | - | + | + | - |
| Jollant et al., 2018 | + | + | + | + |
| Benedetti et al., 2011 | + | + | + | + |
| Canal-Rivero et al., 2020 | + | + | + | + |
| Wagner et al., 2011 | - | + | + | - |
| Besteher et al., 2016 | - | + | + | - |
| Wagner et al., 2012 | - | + | + | - |
| Taylor et al., 2015 | - | + | + | - |
| Cao et al., 2016 | - | + | + | - |
| Tian et al., 2021 | + | + | + | + |
| Cao et al., 2015 | - | + | + | - |
| Fan et al., 2013 | - | + | + | - |
| Wagner et al., 2021 | + | - | + | - |
| Gong et al., 2020 | - | + | + | - |
| Yang et al., 2022 | - | + | + | - |
| Kang et al., 2017 | - | - | + | - |
| Zhang R et al., 2021 | + | - | + | - |
| Cheng et al., 2020 | + | + | + | + |
| Chen et al., 2021 | - | + | + | - |

**Supplementary Table S3. Risk of bias assessment.** + = Low risk of bias; - = Medium risk of bias; x = High risk of bias

|  | **Category 1: Subjects** | **Score** (0/0.5/1) |
| --- | --- | --- |
| 1 | People with a history of suicide attempts were included. Specific diagnostic criteria were applied for the diagnosis of possible mental health conditions. Demographic data was reported |  |
| 2 | People without a history of suicide attempts and overlapping mental health conditions were included and matched to the case cohort. Demographic data was reported |  |
| 3 | Important variables (e.g., age, gender, handedness, height or total brain measures) were checked, either by stratification or statistically |  |
| 4 | Sample size per group > 7 |  |
|  | **Category 2: Methods for image acquisition and analysis** |  |
| 5 | All neuroanatomic measurements were taken without considering group assignment or subject identity |  |
| 6 | Magnet strength > 1T |  |
| 7 | The imaging technique used was clearly described so that it could be reproduced |  |
| 8 | Measurements were clearly described so that they could be reproduced |  |
|  | **Category 3: Results and conclusions** |  |
| 9 | Statistical parameters for significant, and important non-significant, differences were provided |  |
| 10 | Conclusions were consistent with the results obtained and the limitations were discussed |  |
|  | **TOTAL** | /10 |

**Supplementary Table S4. Adapted Imaging Methodology Quality Assessment Checklist**, from

(Cattarinussi et al., 2019), original Checklist available from (Shepherd et al., 2012)

Supplementary results

*Voxel-based Morphometry*

We extracted data for the meta-analysis from nine eligible studies, all published between 2008 and 2020, that used voxel-based morphometry. Two of the included studies ^3,4^ recruited a sample encompassing both people aged under and over 18. Three studies were based in south-east Asia (two in China ^4,5^ and one in South Korea ^6^), one in the USA ^3^ and five in Europe (two in Spain ^7,8^, two in Germany ^9,10^, one in Italy ^11^). The studies conducted in south-east Asia sampled a group of patients with either major depressive disorder (^5,6^ MDD) or a group of mood disorder conditions (^4^, i.e., both bipolar Disorders – BDs – and MDD); the two studies from Spain were conducted with participants with either schizophrenia – SCZ ^7^ or First Episode Psychosis – FEP ^8^. The four remaining studies included patients with MDD ^9,10^ or BDs ^3,11^. Overall, the voxel-based morphometry meta-analysis included 251 people with a lifetime history of SAs and 421 patient controls. The mean age of the participants ranged from 20.5 to 46, with a higher percentage of female participants than males in most studies (ranging from 52,6% to 78.5%), except for the ones on FEP (^8^, 37.7% of females) and SCZ (^7^, 0%). All the significant volumetric differences (number of clusters identified = 66) reported by the studies (*k*=9) included in this CBMA, except for two foci ^11^, identified smaller regional volumes in SA. Nonetheless, no significant spatial overlap between the reported brain regions could be identified at a p < 0.0001 with a cluster-volume of 200 mm^3^ without any statistical correction (and irrespective of considering or not the two studies ^3,4^ whose participants were both over and under 18-years-old). Noteworthily, seven eligible studies ^12–18^ did not find any significant GMV differences between SA and NSA (and thus could not be included in the CBMA).

*Cortical thickness findings*

Six studies that analysed CT differences between people with and without SA were extracted. One of these studies recruited only adolescents ^19^ and was thus excluded from the meta-analysis. Two studies ^20,21^ did not find significant differences between the two groups after correction for multiple comparisons. Thus, three studies were included in an exploratory CBMA. Two studies were conducted in Germany on people with SCZ ^22^ or with MDD ^23^. One study was based in the USA and included people with MDD ^24^. The pooled sample sizes were scarce and consisted of 40 people with a history of SA and 91 without. The mean age ranged from 28.8 to 41; female participants represented 56-57% of the sample (50) to 93.3% ^23^. As for the CBMA based on the brain morphometry, also in this case, no significant spatial overlap between the coordinates of the CT differences could be identified at a p < 0.0001 with a cluster volume of 200 mm^3^ (and without applying any statistical correction for multiple comparisons). Moreover, all the clusters (n= 7) identified by these three studies reported reduced CT in SA

*Task-based fMRI findings*

None of the task-based fMRI studies (*k*=29) retrieved could be compared, as per methodology, to each other, not even when pooling studies that investigated the same cognitive domain with different tasks (i.e., ignoring the tasks employed were different). From the pool of 29 studies, 22 reports were excluded from the CBMA for methodological reasons (see Supplementary Table S2 for reasons for exclusion of each study). Regarding cognitive control tasks, only one study ^25^ met all criteria for inclusion in the CBMA. Nonetheless, no differences between SA and NSA were identified in this study. The study by Vanyukov et al. ^26^ identified several clusters of brain activation abnormalities during risk-taking/delayed reward processing tasks in adults with a history of suicidal behaviour and MDD. However, Potvin and colleagues (76) failed to replicate these findings in patients with SCZ, where a single cluster of reduced prefrontal regions activation was observed. Two studies investigating this cognitive domain could have been meta-analysed yet no differences in brain activation after statistical correction at the whole-brain level were appreciated ^27,28^.

The two eligible studies ^29,30^ investigating emotion processing could not be compared due to methodological reasons (Jollant et al. ^29^ normalised the BOLD signal related to the exposure to angry faces with respect to the signal due to exposure to neutral faces, whereas Ai et al. ^30^ did not).

*Resting-state and task-based FC findings*

Of the studies (*k*=39) investigating FC that could have been considered in a CBMA, only two pairs of studies could be compared, as per methodology, to each other. After excluding those studies that did not meet the inclusion criteria, 23 reports were initially found eligible for further analysis. Two studies ^31,32^ did not identify any differences in FC between the two groups of interest. We divided the remaining twenty-one studies into groups according to the seed region used to compute brain connectivity indexes. Six studies used seeds that were not employed by any other published study that we retrieved, such as the habenula ^33^, the insula ^34^, the posterior cingulate cortex ^35^, the anterior cingulate cortex ^36^, the putamen ^37^ and the ventromedial prefrontal cortex ^38^. Three studies used the ICA approach to study the FC of different networks ^39–41^; one study calculated Global Brain Connectivity (GBC) ^42^, while the two remaining studies parcellated the brain into several ROIs to compute ROI-to-ROI FC indexes (^43,44^; unfortunately, coordinates of the ROIs in ^44^ were not available after contacting the authors. One recent study ^45^ performed FC analysis at the whole-brain level and was thus compared with the study that computed FC based on GBC ^42^. Both studies were conducted in China, with a pooled sample of 45 patients with a mood disorder and a lifetime history of SAs, and 117 patient controls. The mean age ranged between 23 and 35, with a mean percentage of females of approximately 58%. Cheng et al. ^42^ identified one cluster of increased FC in people who attempted suicide (belonging to the left primary auditory cortex), and three clusters of decreased FC (in the right primary sensory cortex, left insula and right visual association area); Chen et al. ^45^ instead identified only two clusters of reduced FC in the right orbitofrontal and right dorsomedial prefrontal cortices. No significant spatial overlap between the coordinates of the clusters identified from the previously cited studies could be identified at a p < 0.0001 with a cluster volume of 200 mm3 (without applying any statistical correction for multiple comparisons).

One study investigated between- and within-network FC differences based on several seeds ^46^. Due to methodological reasons, these studies could not be grouped and meta-analysed together. Two studies from the same group of authors ^47,48^ who applied identical methodologies and analysis pipelines on data from two different populations with past suicidal behaviour did not evince common abnormalities in FC correlated to the dorsal portion of the anterior cingulate cortex.

Five studies used the bilateral amygdalae as seed regions. One of these studies ^3^ was excluded because it investigated the task-based FC, whereas the other four investigated resting-state indexes of FC; two of these studies restricted the indexes of connectivity to be correlated with the amygdalae indexes to either the prefrontal cortex ^49^ or the fronto-limbic circuit ^50^, and thus could not be compared with the other two studies which implemented a whole-brain comparison approach ^51,52^. We conducted an exploratory meta-analysis on the last two studies reported above. Both studies were conducted in South-East Asia (South Korea ^52^ and China ^51^) with a pooled sample of 107 patients with a mood disorder and a lifetime history of SAs compared to 132 patient controls without a history of previous suicidal behaviour. The mean age ranged from 26 to 42, and approximately 74% of the participants from ^51^ were females. Kang et al. ^52^ identified three clusters of altered resting-state FC among SA. Only one of these clusters, belonging to the left medial temporal gyrus, was identified as having abnormal FC with the right amygdala. The other clusters had their FC correlated with the activity of the left amygdala. However, Zhang et al. ^51^ found significant differences only in the FC of two clusters with the right amygdala. Therefore, the two clusters, the activity of which was correlated with the left amygdala (from ^52^), could not be included in the exploratory CBMA.

*Seed-based functional connectivity*

We leveraged the Human Connectome Project Young Adult database to perform a seed-based connectivity analysis to study the functional connections of the right superior temporal gyrus (rSTG). We observed an increase in the FC between the right STG and a bilateral set of brain regions (all p-FWE’s < 0.001 – Figure S1), namely the bilateral precentral gyrus, postcentral gyrus, dorsolateral prefrontal cortex, lingual gyrus, insula, temporal pole, central operculum, lateral occipital cortex, parietal operculum, supplementary motor area and hippocampus. Moreover, we run further corollary correlation analysis from the one reported in the main text: the correlation map of the right STG with left (-36,34,40; p-FWE = 0.00003) and right (32,40,40; p-FWE = 0.000001) DLPFC displayed a positive correlation with friendship scores, while a negative correlation was observed between friendship scores and correlation map of the right STG with the right precentral gyrus/SMA (22,-26,64; p-FWE = 0.000018) and the right postcentral gyrus (16,-18,58; p-FWE = 0.003). Moreover, a positive correlation between the correlation map of the right STG with the right precuneus (0,-66,18; p-FWE = 0.004) and the right DLPFC (26,38,32; p-FWE = 0.03) and self-efficacy scores was found.

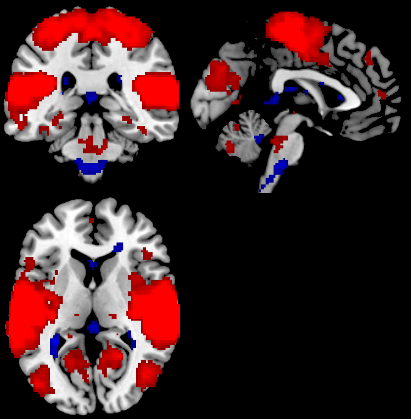

R

**Figure S1. Bilateral set of brain regions with an increase in the FC with the right STG**
